# Deficient hand washing facilities in public toilets in the time of the COVID-19 pandemic: A survey in one high-income country

**DOI:** 10.1101/2021.01.19.21250124

**Authors:** Nick Wilson, George Thomson

## Abstract

**Aims:** To identify the extent of the provision of water and soap for hand washing in public toilets at the time of the COVID-19 pandemic. To also make comparisons with a pre-pandemic survey that included a sample of the same facilities.

**Methods:** We collected data from 400 toilets that were open to the public; all those in three contiguous city council territories (228) and a further convenience sample of 172 around the rest of New Zealand. Comparisons were made with the data on the same facilities included in a 2012/2013 survey.

**Results:** For all the toilets in this survey, 2.5% had no water for hand washing and 14.8% had no soap. There was COVID-19 related health messaging signage in 19.5% of toilets, with posters of the COVID-19 QR code used to facilitate contact tracing in 12.3%, and generic hand washing signage in 1.8%. The hand washing water had “no touch” activation at 28.0% of toilets and at 18.5% for toilet bowl flushing. Toilet bowl lids were not present at 32.8%, and 2.3% of toilets had damage which would impair their functionality (eg, broken toilet seats). For the 128 sites that had also been examined in the previous survey, this new survey found significantly increased provision of soap (risk ratio = 1.47; 95%CI: 1.25 to 1.72), but no increased provision of water.

**Conclusions:** Despite the serious threat of the COVID-19 pandemic, the majority of hand washing facilities in public toilets sampled required tap touching, and did not have health messaging. Nevertheless there has been some modest improvements in soap (but not water) provision since the previous survey eight years before.

## INTRODUCTION

The COVID-19 pandemic has focused international attention on non-pharmaceutical interventions (NPI) to reduce pandemic spread prior to vaccination roll-out. Such NPIs include hygiene practices such as appropriate hand washing, which is an evidence-based measure for preventing respiratory virus transmission.^1^

More specifically, the World Health Organization (WHO) has issued guidance on COVID-19 and hygiene/sanitation.^2^ This guidance highlights the importance of hand washing facilities (with water and soap) and of having separate toilets “for people with suspected or confirmed SARS-CoV-2 infection.” Also “the toilet should be flushed with the lid down to prevent droplet splatter and aerosol clouds.” The latter recommendation is supported by data from a COVID-19 quarantine room study which found that “the inner walls of toilet bowl and sewer inlet were the most contaminated sites with the highest viral loads.”^3^ Another such study reported that “there was extensive environmental contamination by 1 SARS-CoV-2 patient” with toilet bowl and sink samples being positive for viral RNA.^4^ There has also been one study indicating circumstantial evidence of fecal aerosol transmission of COVID-19 via an apartment drainage system,^5^ similar to an outbreak from fecal aerosols of SARS-CoV-1 in 2003.^6^

While improving hygiene in a pandemic is highly desirable, it is also beneficially for reducing the spread of other respiratory viruses such as seasonal influenza and norovirus infection. For example, one review identified six studies that implicated bathroom surfaces as primary sources of human norovirus infection.^7^

In our case study country, New Zealand, there was community spread of the pandemic virus (SARS-CoV-2) on a number of occasions during the 2020 year, but elimination was successfully achieved and re-achieved after a series of border control failures.^8 9^ This success arose largely from a combination of tight border controls (quarantine and isolation), a strict lockdown and widespread testing/contact tracing. Actions related to hygiene included:

- Mass media messages relating to hand washing, cough etiquette, staying home when sick, and mask use.
- Actions by organisations to install hand sanitisers and posters with hygiene messaging in workplaces and various public places.
- Public toilets were all closed at the highest lockdown level.
- Actions by some local government agencies to increase soap availability in the public toilets they maintain (eg, by Napier City Council^10^) and to include posters with hygiene messaging in these facilities.

Given the latter point, the aim of this study was to identify the provision of water and soap for hand washing (along with related health messaging) in New Zealand public toilets at the time of the COVID-19 pandemic. We also aimed to make comparisons with a pre-pandemic survey^11^ that included a sample of the same facilities.

By way of context, New Zealand is a high-income country, but one where there has been reported shortages of public toilets, especially in areas with high numbers of international tourists.^12 13^ Previous research has also shown deficiencies with public toilets in the country, in terms of lacking hand washing water (4%) and lacking soap (39%).^11 14^ Another study of toilets in one New Zealand city (mainly at cafés and public facilities), found that some had no handwashing facilities (2%) and no soap (13%).^15^ Within this sample, the lack of soap was highest in the public toilets at 38%. Another New Zealand study of primary school toilets also reported that only 28% had facilities meeting the relevant code of practice ie, there was a lack of hot water, lack of drying facilities and lack of soap.^16^ Inadequate hand hygiene has also been recorded, with public toilet users in New Zealand sometimes not washing hands (13%) or using soap (28%).^17^

## METHODS

### Survey sampling

The sample frame was comprehensive for the public toilets in three contiguous cities with a combined population of 389,000, with additional convenience sampling in cities and rural areas, based on author travel plans for other reasons around New Zealand from 18 July 2020 to 2 January 2021. We expanded the sample to a total of 400 toilets, which we estimated would be at least 10% of the facilities nationwide. But we also aimed to replicate a previous survey of facilities in public toilets in the lower North Island in 2012/2013.^11^ The public toilets were located by:

- Online city council maps of public toilets were used for the comprehensive three council area survey.^18-20^ However, data could not be collected on a minority of these (<5%) due to closure for repairs and/or being padlocked shut, suggestive that they were only opened for specific sporting events.
- Using ‘Google Maps’ to locate a ‘city/town/district’, and then in the map searching for ‘public toilets’.
- Using the smartphone app version of ‘CamperMate’.^21^
- Roadside public toilet signs when travelling by car.

We excluded from the sample, public toilets that were not directly open to the outside (ie, which were inside of other buildings such as within shopping complexes; council-owned facilities such as libraries; and within railway or ferry buildings, some of which were signed as for “patrons only”). Where facilities were closed or were being cleaned, we attempted repeat visits where this was convenient.

### Data collection

At each toilet facility, we surveyed all the men’s and unisex toilets. Toilet facilities contained from one to six self-contained “toilet units” with a separate door that opened to the outside – albeit some units of “men’s toilets” would include multiple hand washing basins, a urinal and multiple toilet bowls. Data were collected on the availability of water and soap for handwashing. Toilets with empty soap containers were counted as without soap. We also photographed all health-related signage (eg, relating to COVID-19 and handwashing). Requirements for touching (or not) of taps/buttons/levers for activating tap water and flushing the toilet were noted. The presence of lids for toilet bowls was documented, given concerns around potential virus dispersion when flushing when the lid is not down (see *Introduction*).

### Analysis

We compared results for the three city council comprehensive sample with the supplementary convenience sample. Comparisons were also made with the exact same facilities involved in the previous survey conducted in 2012/2013 (albeit excluding 14.7% (22/150) which were not readily accessible or which had subsequently been closed down). Statistical analysis used OpenEpi (v 3.01) and Mid-P exact values were calculated (2-tailed).

### National denominator estimation

To provide context for our survey, we estimated the total number of such public toilet facilities in New Zealand using the following steps:

- For city-based local authorities we used as a basis the facilities we surveyed in the three city comprehensive sample (n=131 or 3.4 per 10,000 population).
- For the other local authorities we used data from Google Maps in three LAs in the Wairarapa region (n=13 facilities) and scaled from our estimate of the sensitivity of such data on Google Maps from the three city council survey (at 61.8% or 81/131 giving a scaling factor of 1.62). This gave 21 facilities or 4.3 per 10,000 population.
- We then extrapolated the three city council results to the other 12 LAs in New Zealand which were city ones; and the Wairarapa results to the other 51 LAs which were council districts.

## RESULTS

### Survey results for 2020/21

Our survey comprised 400 toilet units at 242 toilet facilities. We estimated that nationwide the total number of such toilet facilities was around 985 in city councils and around 760 in district councils, ie, around 1864 in the whole country (3.7 per 10,000 population). So our sample was estimated at 13.0% of the estimated total (242/1864), which was greater than our target of a 10% sample.

Most of the sample of 400 toilet units were from the comprehensive survey of three contiguous city councils (n = 228), relative to the additional convenience sample (n = 172). The former grouping was all in the lower North Island, were more likely to be unisex (vs men’s toilets), and involved more toilets in cities (but fewer in towns and rural areas) (Table 1).

**Table 1:** Full results for the 2020/2021 survey of public toilets in New Zealand (n=400 units surveyed; n=242 toilet facilities, showing column percentages)

| Characteristic | Comprehensive sample of 3 city LAs (n=228 units unless indicated otherwise) |  | Additional convenience sample (n=172 units unless indicated otherwise) |  | Total (n=400 toilet units) |  | Comments |
| --- | --- | --- | --- | --- | --- | --- | --- |
|  | N | % | N | % | N | % |  |
| **Type, location** |  |  |  |  |  |  |  |
| Facilities (ie, toilet blocks, some with multiple toilet units that opened to the outside) | 131 | – | 111 | – | 242 | – | That is the average toilet facility had 1.7 separate toilet units (400/242) (median=1 unit; range: 1 to 6 units). All but one facility had no user charges; and one facility had an office with a supervisor. |
| Men's | 44 | 19.3% | 65 | 37.8% | 109 | 27.3% | Four were urinals only. |
| Unisex | 184 | 80.7% | 107 | 62.2% | 291 | 72.8% |  |
| In the North Island | 228 | 100.0% | 126 | 73.3% | 354 | 88.5% | The range was from Auckland to Wellington |
| In the South Island | 0 | 0.0% | 46 | 26.7% | 46 | 11.5% | The range was from Picton to Christchurch |
| In a city | 223 | 97.8% | 29 | 16.9% | 252 | 63.0% | In NZ a city is defined in the Local Government Act (2002) as having a population of over 50,000. This meant the following cities were included: Auckland, Christchurch, Hutt, Napier, Palmerston North, and Porirua. |
| In a town | 5 | 2.2% | 126 | 73.6% | 131 | 32.8% | A town was defined as any settlement with a population below that of a city. |
| In a rural area | 0 | 0.0% | 17 | 9.9% | 17 | 4.3% |  |
| **Water for hand washing** |  |  |  |  |  |  |  |
| Water not available | 1 | 0.4% | 9 | 5.2% | 10 | 2.5% |  |
| Automatic, no-touch water delivery | 104 | 45.6% | 8 | 4.7% | 112 | 28.0% |  |
| – Lever mechanism for tap | 5 | 2.2% | 11 | 6.4% | 16 | 4.0% | This is a subset of the above row. |
| **Soap** |  |  |  |  |  |  |  |
| Not available | 22 | 9.6% | 37 | 21.5% | 59 | 14.8% |  |
| – Dispenser not working / empty | 12 | 5.3% | 3 | 1.7% | 15 | 3.8% | This is a subset of the above row. |
| Bar/cake soap only | 0 | 0.0% | 24 | 14.0% | 24 | 6.0% |  |
| **Toilet bowls** |  |  |  |  |  |  |  |
| Automatic flushing (no need to use a button or lever) | 58/226 | 25.7% | 14/163 | 8.6% | 72/389 | 18.5% | Denominator was facilities (excluding urinal only facilities) |
| Lid missing | 78/240 | 32.5% | 68/205 | 33.2% | 146/445 | 32.8% | Denominator includes all separate toilet bowls |
| **Urinal flushing** |  |  |  |  |  |  |  |
| Automatic flushing | 52/55 | 94.5% | 43/63 | 68.3% | 95/118 | 80.5% | Other urinals required a button/level/cord to be used |
| **Notable facility damage** |  |  |  |  |  |  |  |
| Damage | 6 | 2.6% | 3 | 1.7% | 9 | 2.3% | See footnote.* |
| **Health-related signage** |  |  |  |  |  |  |  |
| Any COVID-19 related behavioural messaging | 33 | 14.5% | 45 | 26.2% | 78 | 19.5% | Excluding QR codes – see below. There was an example of a hand washing sign involving soap in a toilet with no soap available. |
| Any COVID-19 QR code signage | 27 | 11.8% | 22 | 12.8% | 49 | 12.3% | Inside or on outside wall/door. We included one sign that had fallen onto the floor. |
| Any hand washing signage | 7 | 3.1% | 0 | 0.0% | 7 | 1.8% | That is generic signage, not COVID-19 specific |
| Any non-smoking signage | 2 | 0.9% | 3 | 1.7% | 5 | 1.3% |  |
| No health-related signage | 175 | 76.8% | 116 | 67.4% | 291 | 72.8% | None of the above 4 categories |

\* Damage included broken seats, a toilet bowl lid with a hole, soap dispenser container remnants, the light not working, and a toilet roll holder on the ground.

The proportion of all the 400 toilets without water for hand washing was 2.5%, with this being higher in the convenience sample than the three urban councils sample (5.2% vs 0.4%; risk ratio (RR) = 11.9; 95%CI = 1.53 to 93.3; p = 0.0030). Absence of soap was 14.8% overall, and this was also higher in the convenience sample vs the three councils sample (21.5% vs 9.6%; RR = 2.23; 95%CI = 1.37 to 3.64; p = 0.0011). Most soap was dispensed as a liquid or foam (Figure S1-3), but at 6.0% of toilets it was available in a cake/bar form. A number of toilets had containers for liquid soap, but they were empty (Table 1).

In terms of “no touch” activation, this was available for hand washing water at 28.0%, for toilet bowl flushing at 18.5%, and for urinal flushing at 80.5%. Toilet bowl lids were not present for 32.8% (many were designed or built this way) (Figure S3-1), and 2.3% of toilets had damage which would impair their functionality (eg, broken toilet seats, Figure S3-1; broken toilet rolls, Figure S3-2; and destroyed liquid soap dispensers, Figure S3-3).

The majority of toilets had no health related-signage (72.8%). Some form of COVID-19 related health messaging was the most common type (19.5%), followed by the COVID-19 QR code used to facilitate contact tracing via an official smartphone app (12.3%), generic hand washing signage (1.8%) and then non-smoking signage (1.3%), (see Supplementary File 2 for examples of these posters). COVID-19 health messaging signage was more common in the convenience sample than in the three urban council sample (RR = 1.81; 95%CI = 1.21 to 2.71; p = 0.0040). There was no signage that promoted toilet lid lowering prior to flushing.

From a qualitative perspective, we noted that several COVID-19 signs in tourist areas were in Chinese language (Figure S2-1) and a few hand washing signs included te reo Māori wording (language of Indigenous New Zealanders) (Figure S2-3). We also noted that some automatic water and soap dispensers took some time to activate and then dispensed too little soap or too little water for a satisfactory hand wash (ie, repeat activation was required).

### Survey results for 2020/21

The comparison of the exact same toilet units involved in the previous survey in 2012/13 is shown in Table 2. There was no improvement in the availability of water for hand washing, but soap availability improved significantly from 59% to 86% (risk ratio = 1.47; 95%CI: 1.25 to 1.72).

**Table 2:** Comparison of the exact same public toilet units in the two surveys (albeit with minor differences in numbers due to some being demolished or closed at the time of the second survey*; all facilities were in the lower North Island)

| Characteristic | Previous survey in 2012/13<sup>11</sup> |  | This survey in 2020/21 |  | P-value |
| --- | --- | --- | --- | --- | --- |
|  | N | % | N | % |  |
| Water available for hand washing | 123/128 | 96.1% | 123/128 | 96.1% | 1.0 |
| Soap available** | 75/128 | 58.6% | 110/128 | 85.9% | <0.0000001 |

\* If a facility had changed in the number of toilets (eg, expanded from 2 to 3 units); we only compared the exact same numbers as in the original survey.
\*\* In liquid, foam or bar/cake forms.

## DISCUSSION

### Main findings

This survey found a deficient provision of soap (14.8% of toilet units with none), and to a lesser extent water (2.5% with none). These findings suggest that while there has been some statistically significant improvement in soap provision (but not water provision) in the eight year period since the previous survey, the attempts by some local government agencies to increase soap provision at the time of the COVID-19 pandemic^10^ needs to be further augmented.

The higher water (5.2% vs 0.4%) and soap provision (21.5% vs 9.6%) in the comprehensive three urban council sample vs the convenience sample respectively, is likely to reflect the more modern facilities in the former and/or a higher quality maintenance schedule. As New Zealand is highly urbanised, we suspect that the true nationwide results would be closer to those of the three council survey than the convenience sample ones.

The relatively low level of health related signage was a problematic finding, especially the COVID-19 related health messaging (at 19.5%), the COVID-19 QR code (12.3%), and generic hand washing signage (1.8%). These deficits are likely to represent both long-term lack of signage relating to hand washing, as well as an inadequate response to the COVID-19 threat.

Also of note from a hygiene perspective was the limited extent of “no touch” activation of devices (ie, for hand washing water at 28.0% and for toilet bowl flushing at 18.5%). These should ideally be expanded with the potential long-term goal being to have maximally “no touch” facilities (including automated: door opening and locking via hand waving in front of a sensor, water dispensing, soap dispensing, toilet flushing and hand drier activation). Automation could extend to the toilet bowl lid being closed before flushing.

Ensuring that all toilet bowls have lids (missing for 32.8% in this survey – often by design) is also desirable, along with messaging to close the lids prior to flushing (see *Introduction* for the rationale for lid closing).

### Study strengths and limitations

This is the largest such survey to date in New Zealand that we know of, and it was able to compare a sub-sample of the same facilities after an eight year period. It was also conducted at a time where there was greatly heightened need for hygiene due to the COVID-19 pandemic.

Nevertheless, the study was limited by only being comprehensive in three contiguous council areas, with the rest being convenience sampling. This was owing to this being an unfunded study with no budget for travel. The convenience sampling is likely to have involved surveying facilities that were more accessible to the researchers by being on main roads. These may be relatively newer and have a better maintenance schedule than facilities in small rural towns. This may have resulted in some under-estimation from the convenience sample of the extent of the problems outside urban areas (eg, with water and soap availability).

### Potential research and policy implications

A fundamental research issue is to better quantify the risks of infectious disease transmission associated with use of public toilets (eg, from any aerosolisation of faeces and from touching contaminated surfaces). This is not only relevant to SARS-CoV-2, but also other infectious diseases (eg, norovirus infection and seasonal influenza).

Nevertheless, surveys such as this could be improved upon by making them fully random at the national level and collecting additional data on the facilities to compare them with local standards (eg, as per New Zealand ones,^22^ albeit from 1999) or more state-of-the-art Japanese designs.^23^ Research on ways to minimise the vandalism of public toilets is also needed, as this has been a reported barrier to soap dispensers in New Zealand in the past.^10^ Art work inside toilets and on exterior walls is used in some New Zealand toilets (Figure S1-1) and might be worth expanding if it is found to be effective against vandalism.

Policy goals for local government could be to ensure all facilities have water and soap, and to move towards designs that are maximally “no touch”. Built-in redundancy (eg, two separate soap dispensers) may be desirable to minimise the risk of running out of soap, and cakes of soap should probably be avoided as these are more vulnerable to theft. If fully automated taps are not installed, then tap levers or floor pedals for activating water flow could be an alternative. Levers allow users to use the back of their hands and levers can also be more suited than conventional taps for people with disabilities such as arthritis.

Central government could set and enforce minimum standards for council-owned public toilet facilities and the extent of their provision. It could also boost funding support for these purposes, at least partly from border charges collected from international tourists (a funding system already in place in New Zealand). The extent of the funding needed should be seen in the context of the huge costs of COVID-19 and future pandemics, and from other respiratory viruses such as seasonal influenza and from norovirus infection.

## Conclusions

To conclude, despite the serious threat and great costs of the COVID-19 pandemic, and while there has been some improvement in soap provision in the eight year period since the previous survey, the attempts by some local government agencies to increase such provision need to be further augmented. There are also other design and maintenance deficiencies that would improve hygiene in public toilets. There is a major scope for improving health messaging at these sites, and this might be a low-cost and quick intervention to reduce pandemic spread.

## Data Availability

The authors confirm that the data supporting the findings of this study are available within the article.

