## Supplementary file 1 for "Deficient hand washing facilities in public toilets in the time of the COVID-19 pandemic: A survey in one high-income country"

Figure S1-1: Exterior art work on a public toilet which may reduce the risk of graffiti attacks and vandalism (toilet in the suburb of Maungaraki, Lower Hutt) (photograph by first author, 2020)

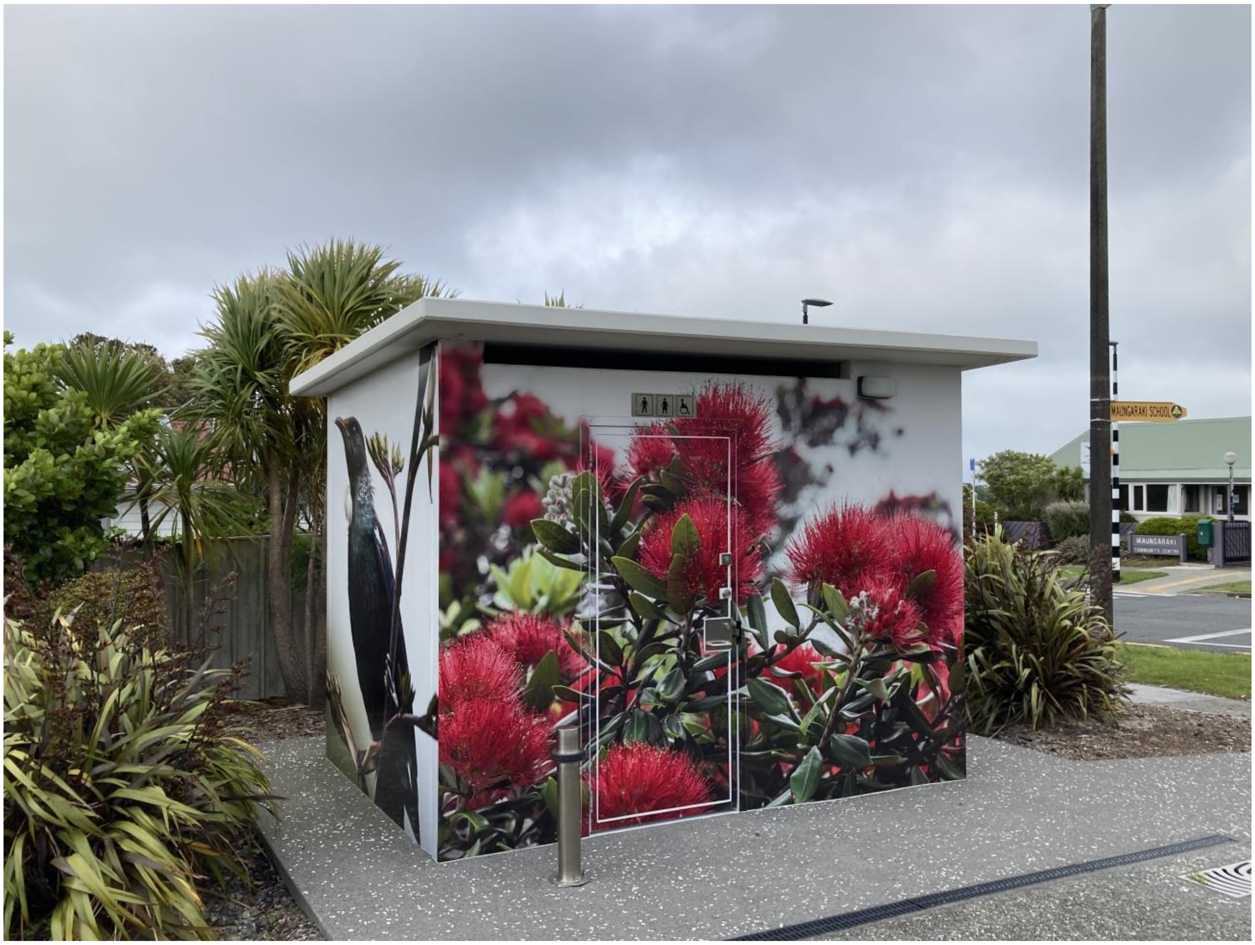

Figure S1-2: Dilapidated entrance to a public toilet with no signage as to it being men's or women's on Island Bay Beach Wellington (photograph by first author, 2020)

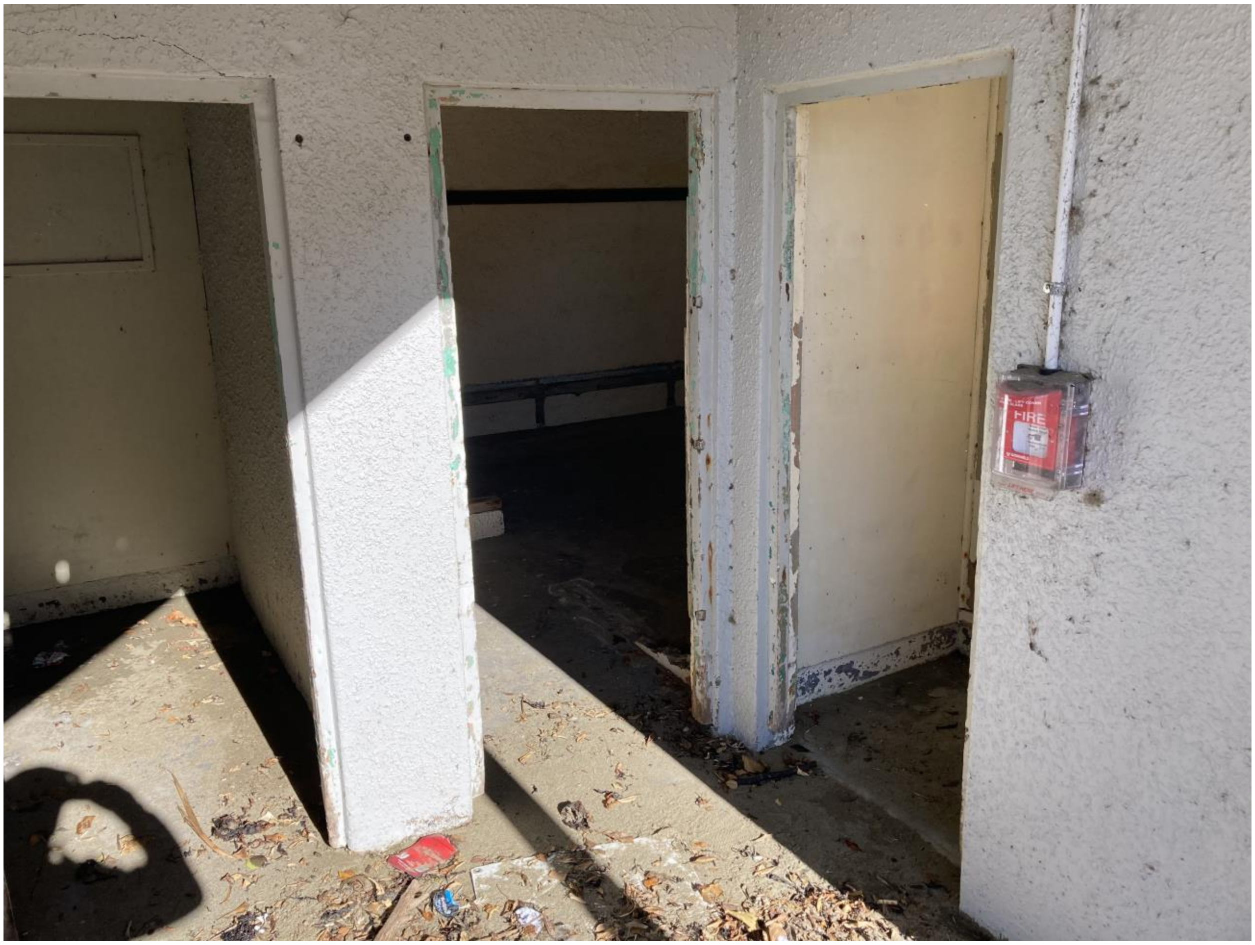

Figure S1-3: The most common type of toilet arrangements found in this 2020/2021 study ie, manual tap, liquid soap dispenser, manual flushing and a lid over the toilet bowl (photograph by first author, 2020)

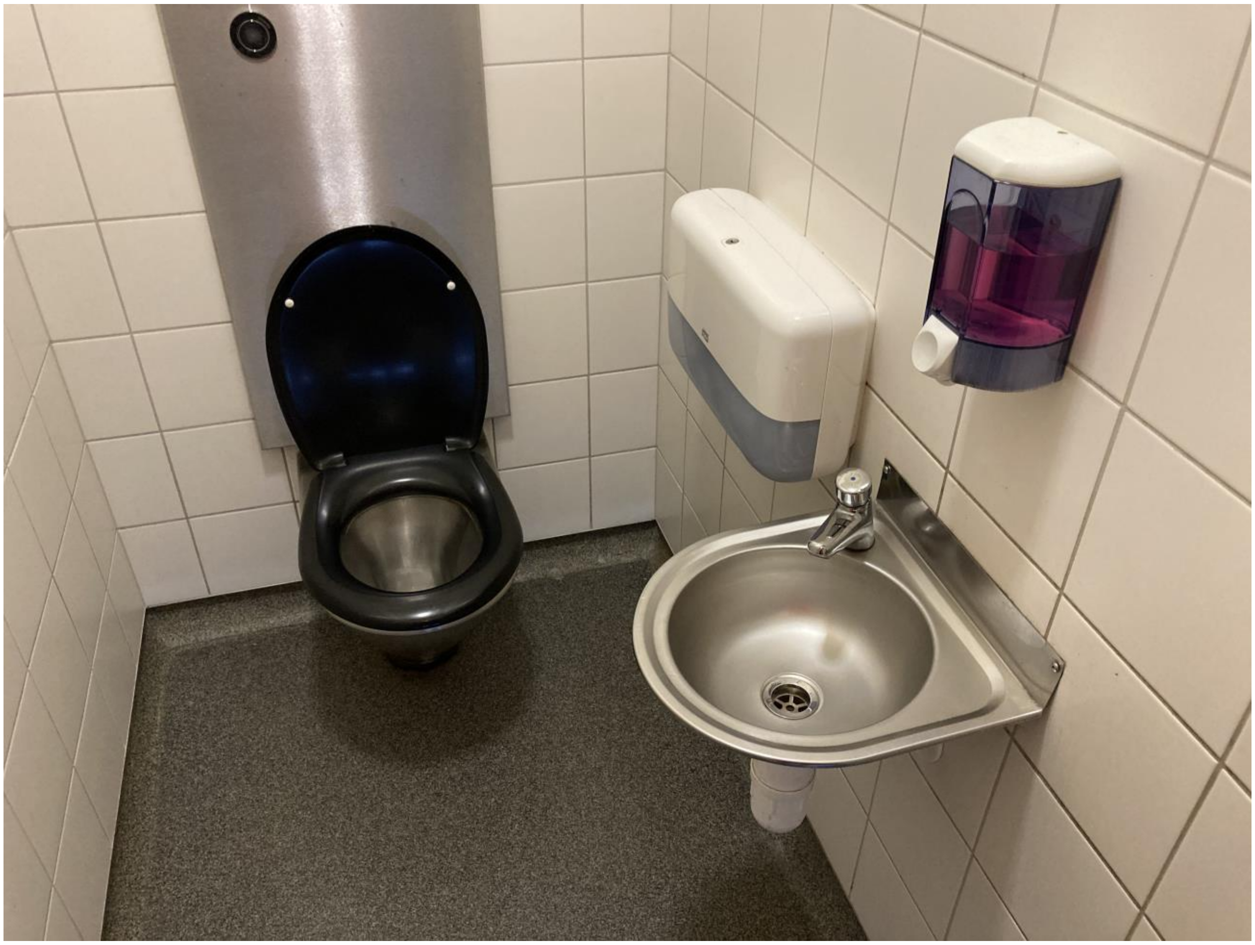
