## Supplementary file 2 for "Deficient hand washing facilities in public toilets in the time of the COVID-19 pandemic: A survey in one high-income country"

Figure S2-1: A relatively common type of COVID-19 hygiene poster on a toilet wall on the right, with the Chinese language equivalent on the left (photograph by first author, 2020)

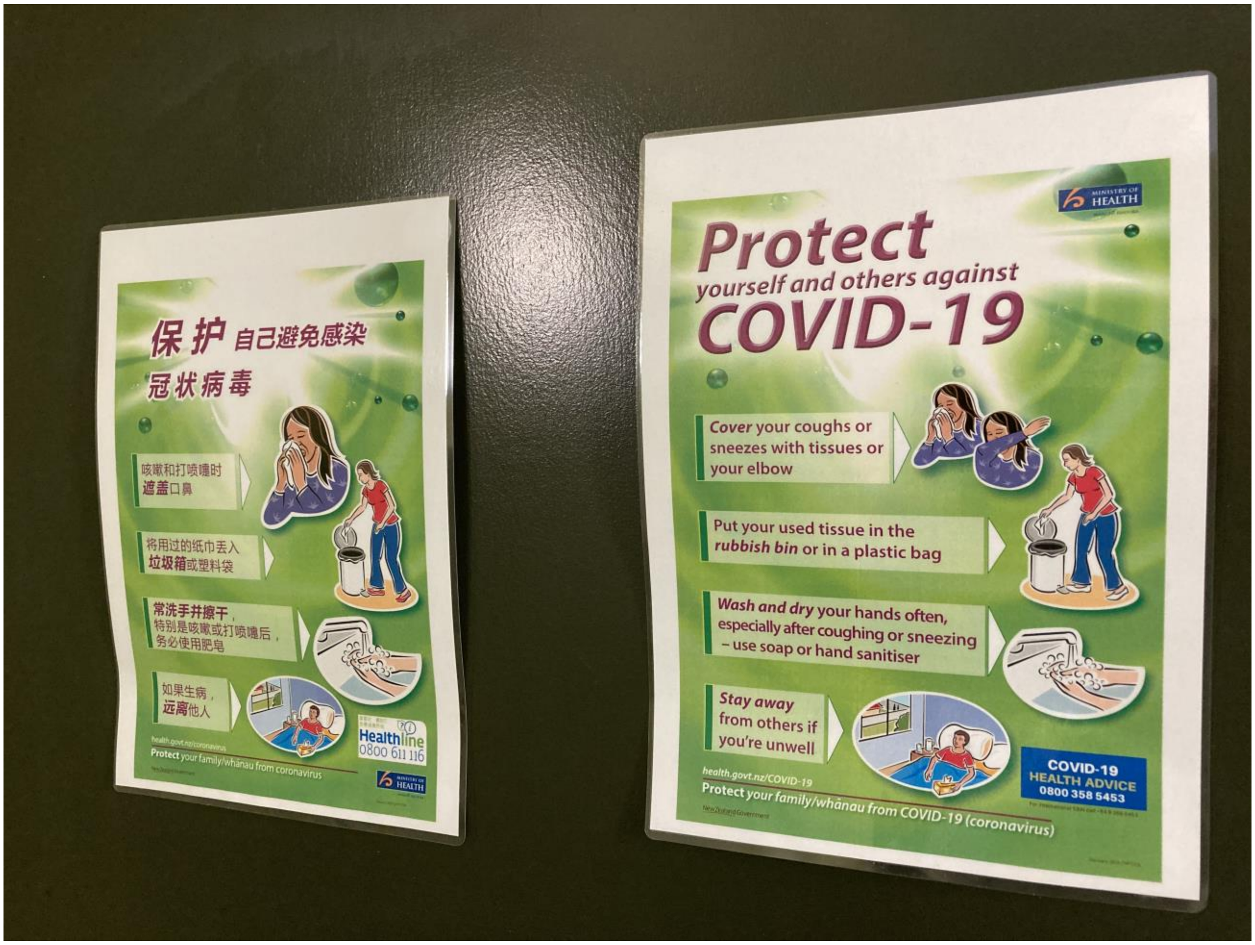

Figure S2-2: A partly vandalised COVID-19 poster inside a toilet changed to “unite against government” (photograph by first author, 2020)

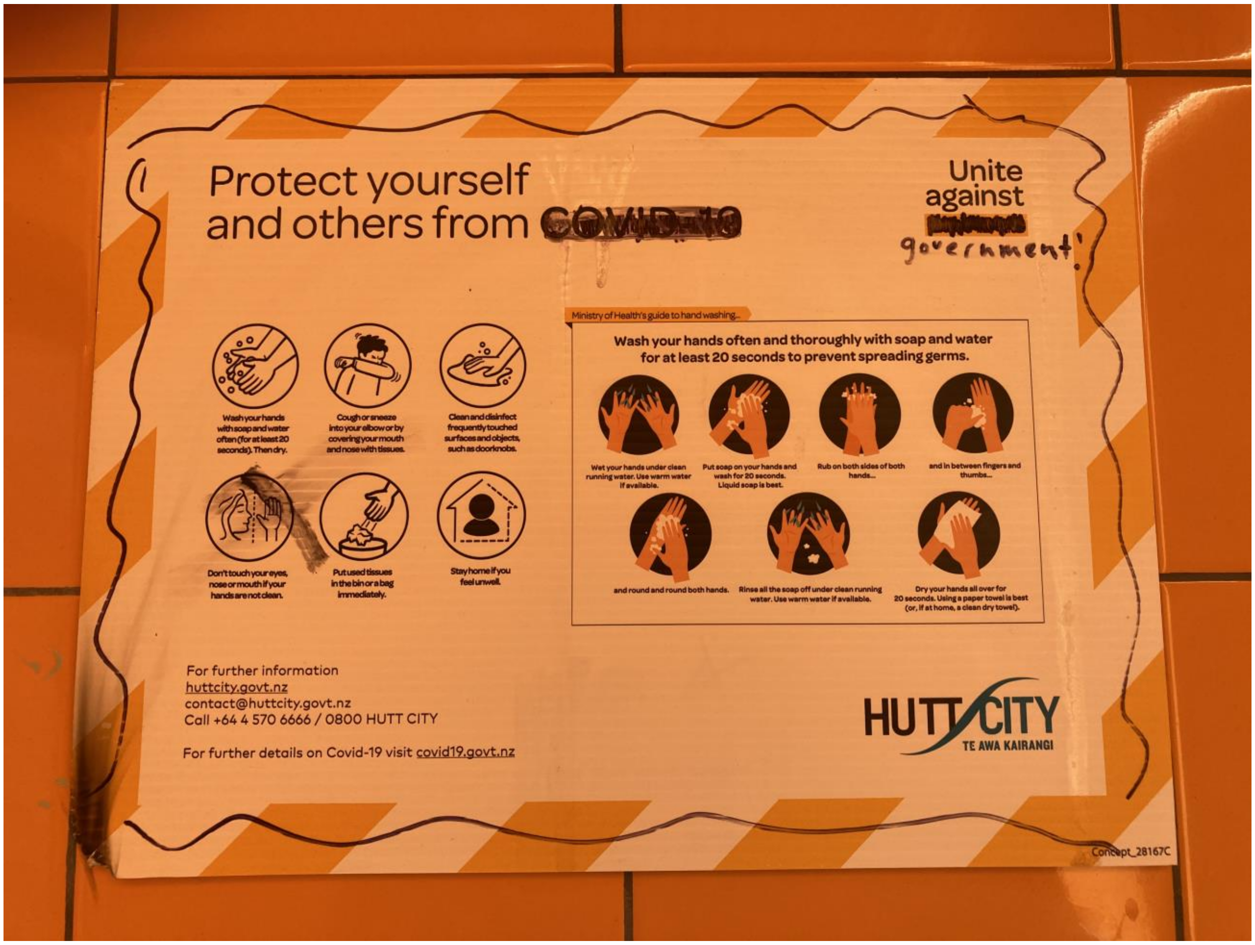


Figure S2-3: A rare “wash your hands” sign that has wording in te reo Māori, the language of Indigenous New Zealanders (photograph by first author, 2021)

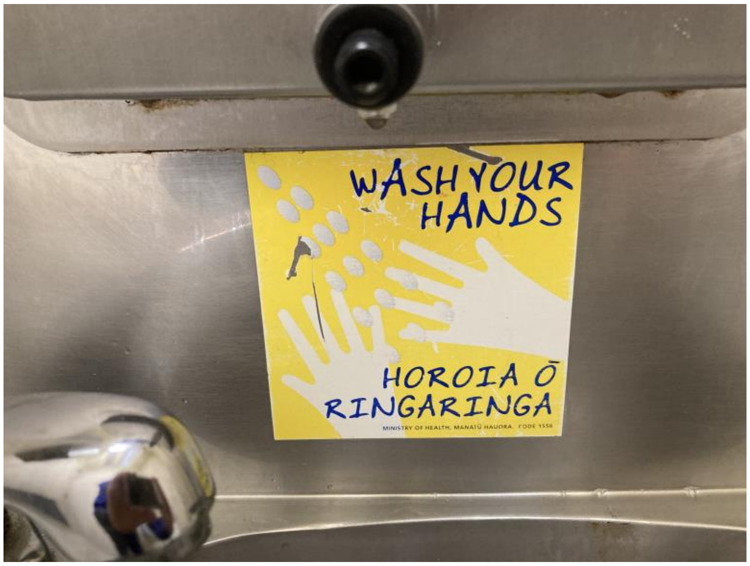

Figure S2-4: On the outside of a unisex toilet door, a QR code for scanning with the NZ COVID Tracer smartphone app to facilitate contact tracing (photograph by first author, 2020)

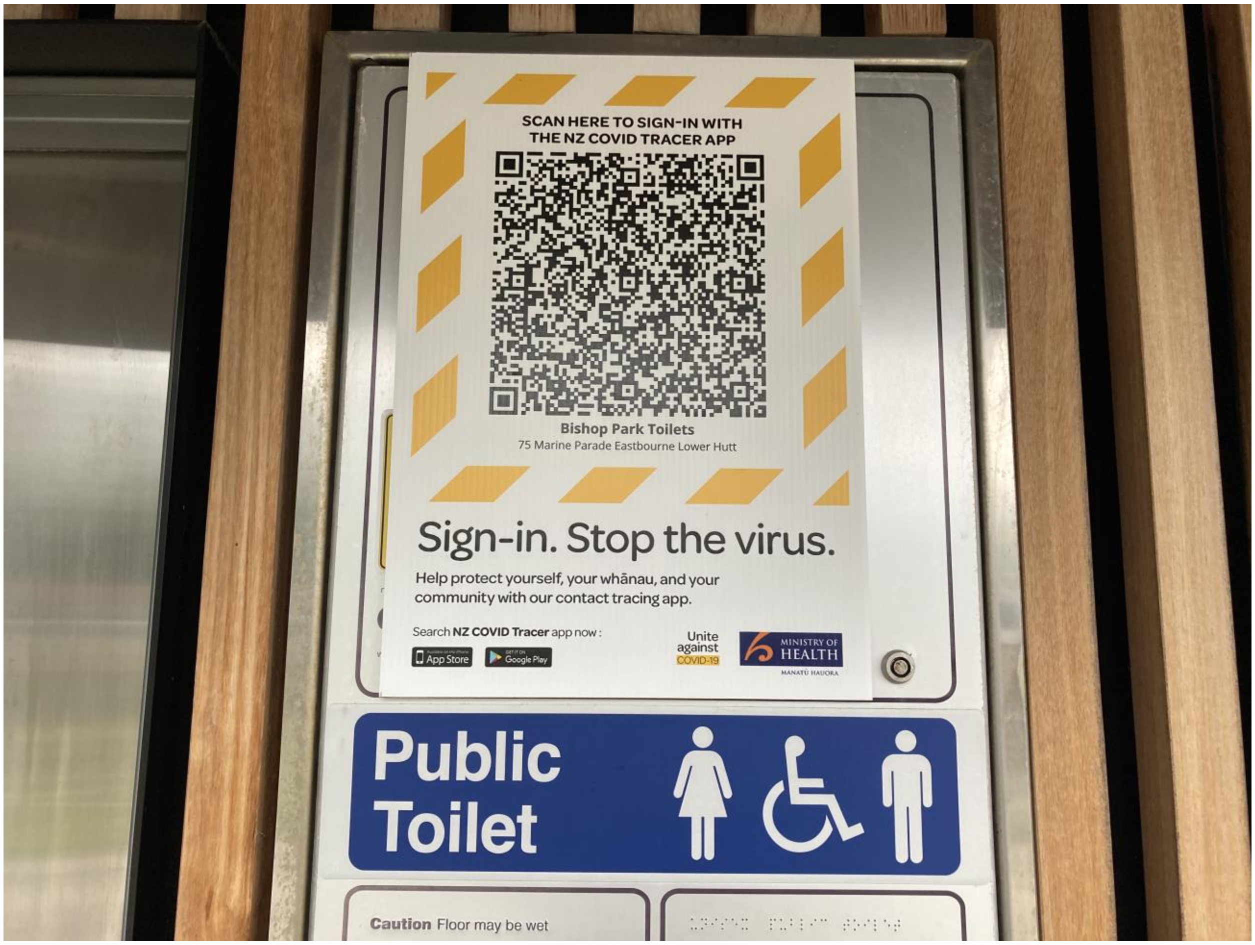

Figure S2-5: COVID-19 posters used in public toilets (photograph by first author, 2020)

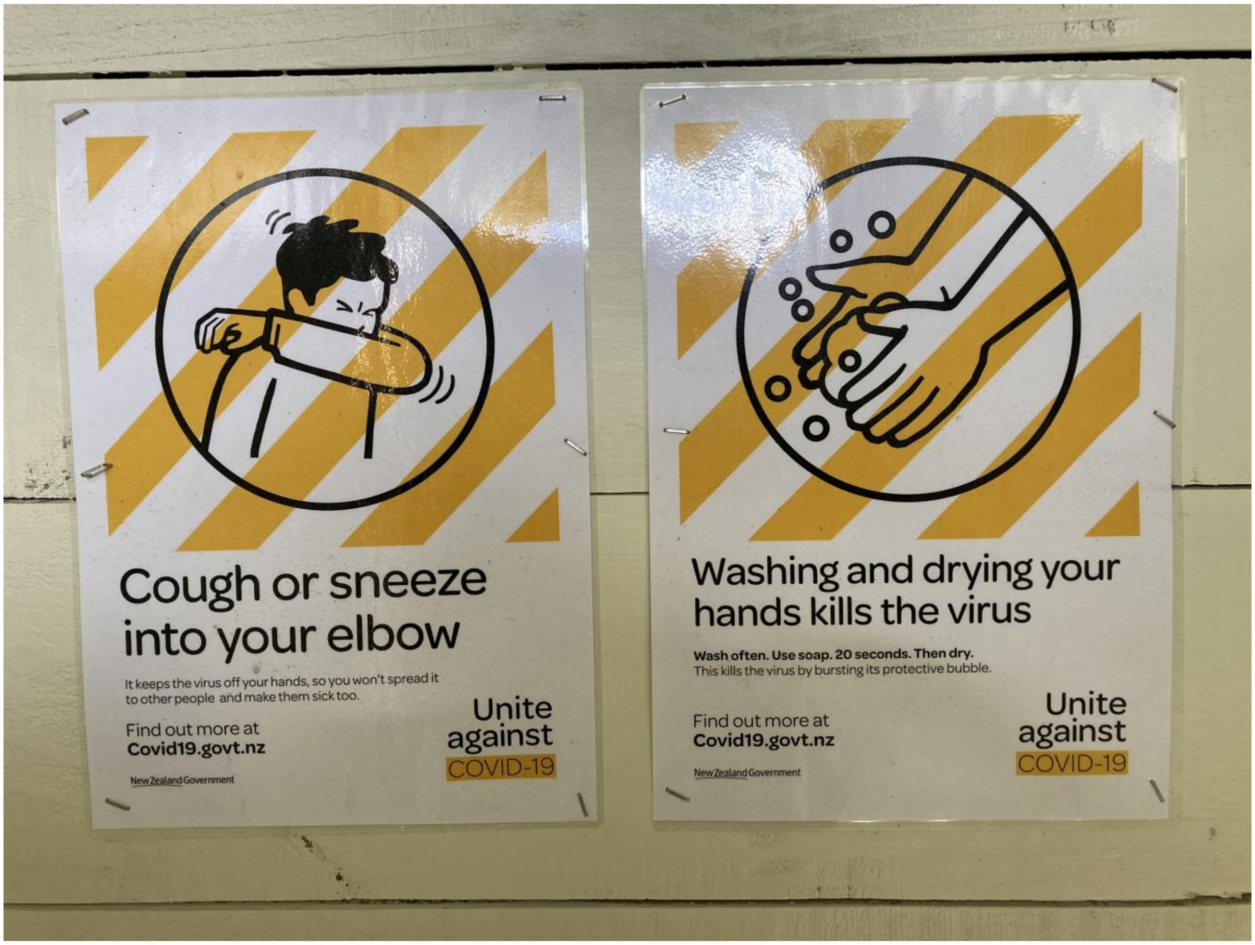
