## Supplementary file 3 for "Deficient hand washing facilities in public toilets in the time of the COVID-19 pandemic: A survey in one high-income country"

Figure S3-1: Example of a broken toilet seat which also has no lid. This photo also shows a lever for activating the tap and a cake of soap (photograph by first author, 2021)

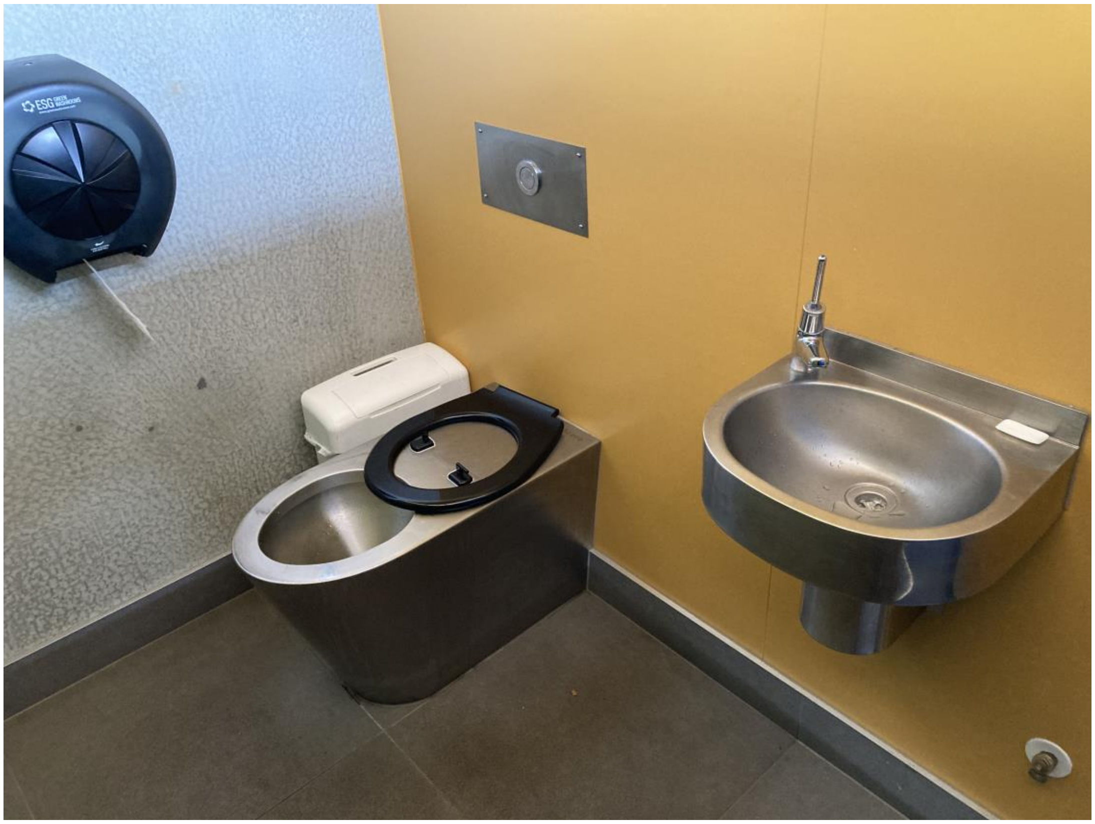

Figure S3-2: Example of a toilet with a broken toilet paper dispenser (photograph by first author, 2021)

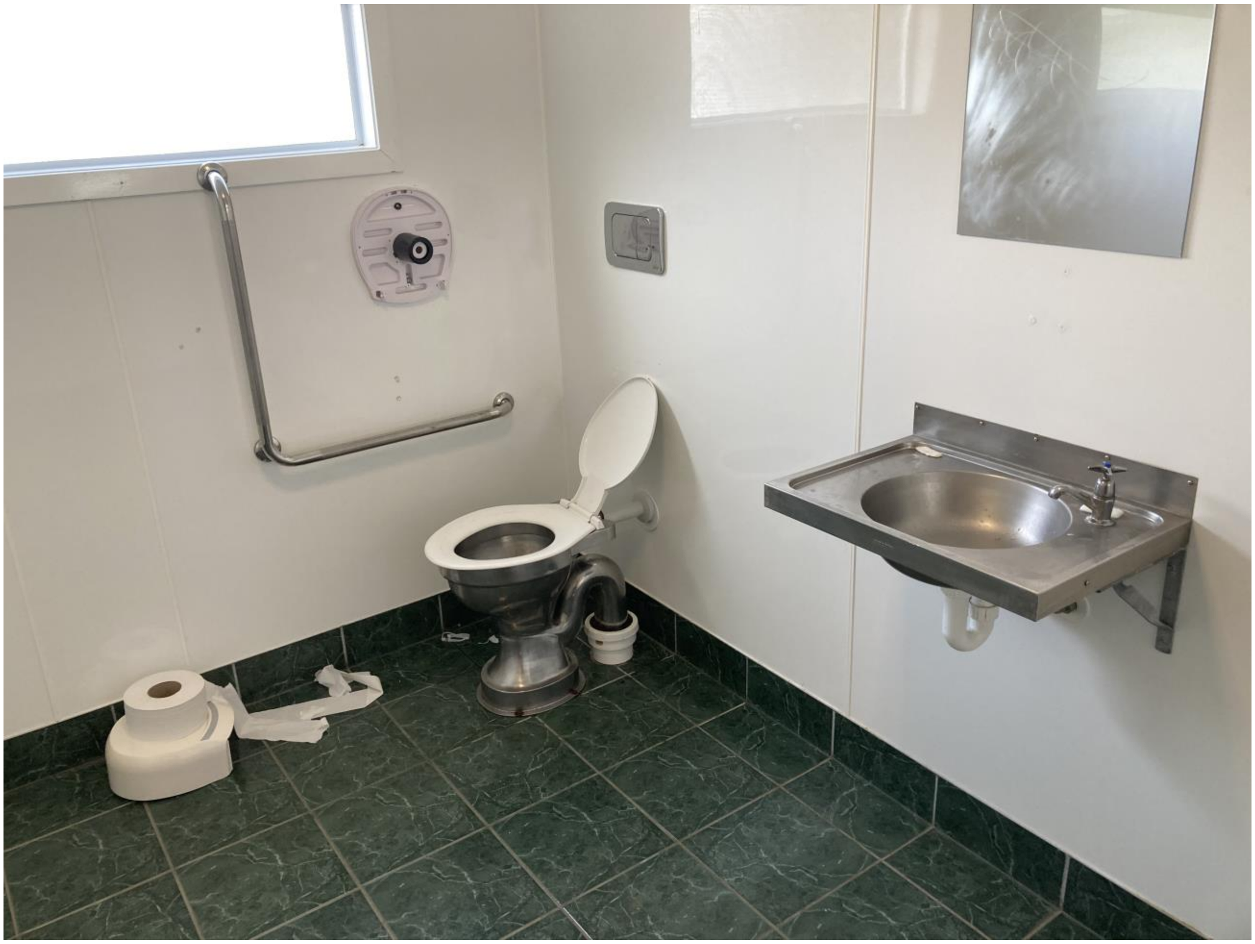

Figure S3-2: Example of a destroyed liquid soap dispenser below (photograph by first author, 2020)

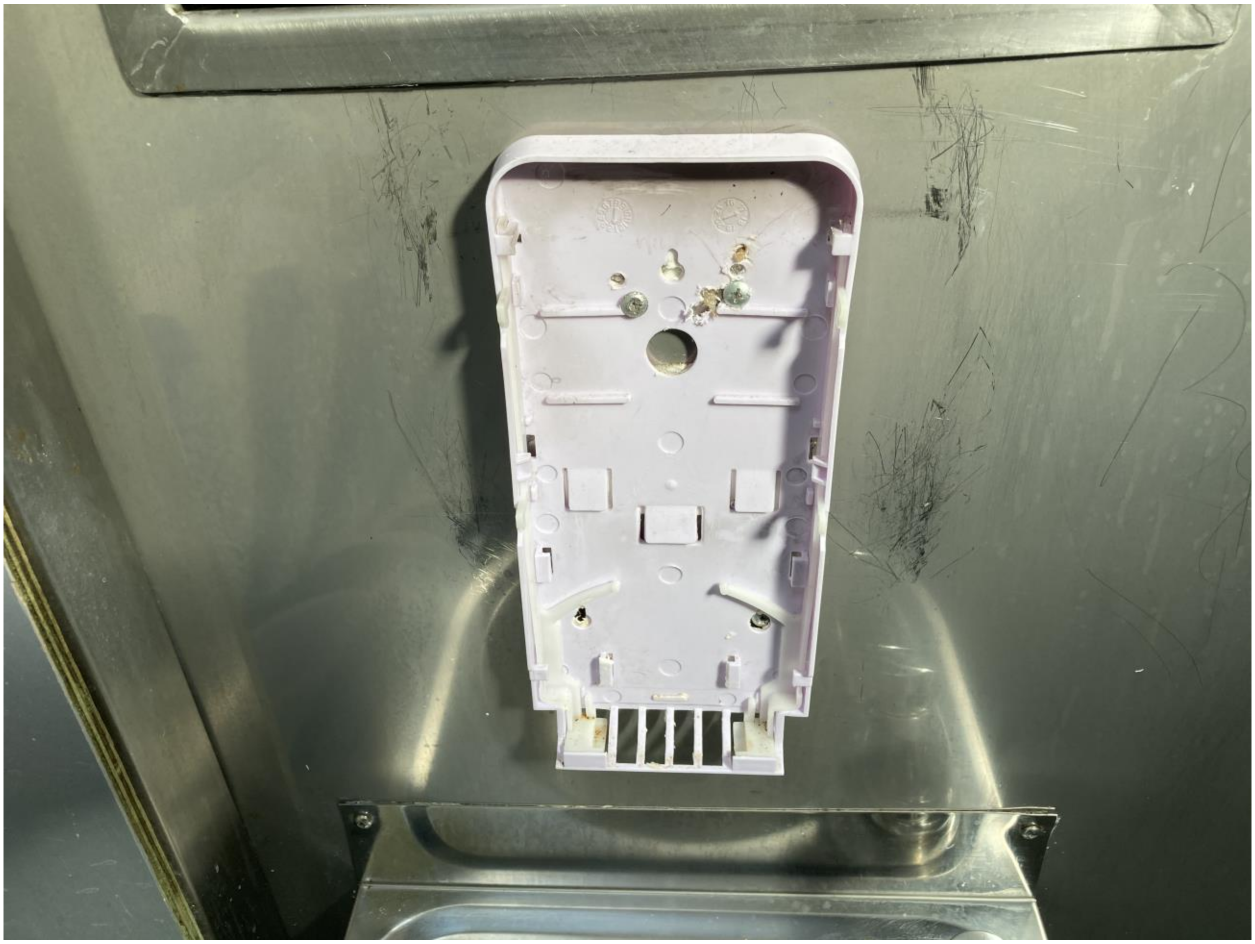
